# The clinical phenotype of autism spectrum disorder in individuals with 3q29 deletion syndrome

**DOI:** 10.1101/2022.11.01.22281767

**Authors:** Rebecca M Pollak, Jordan E Pincus, T Lindsey Burrell, Joseph F Cubells, Cheryl Klaiman, Melissa M Murphy, Celine A Saulnier, Elaine F Walker, Stormi Pulver White, Jennifer G Mulle

**Affiliations:** Center for Advanced Biotechnology and Medicine, Robert Wood Johnson Medical School, Rutgers University; Department of Pediatrics, School of Medicine, Emory University; Marcus Autism Center, Children’s Healthcare of Atlanta and Emory University School of Medicine; Clinical Psychology, College of Arts & Sciences, Georgia State University; Department of Human Genetics, School of Medicine, Emory University; Department of Psychiatry and Behavioral Science, School of Medicine, Emory University; Neurodevelopmental Assessment & Consulting Services; Department of Psychology, Emory University; Department of Psychiatry, Robert Wood Johnson Medical School, Rutgers University

**Keywords:** 3q29 deletion, autism, copy number variants, developmental delay, genomic disorder, psychiatric genetics, ADOS-2, ADI-R

## Abstract

**Background:** The 1.6 Mb 3q29 deletion is associated with neurodevelopmental and neuropsychiatric phenotypes, including a 19-fold increased risk for autism spectrum disorder (ASD). Previous work by our team identified elevated social disability in this population via parent-report questionnaires. However, clinical features of ASD in this population have not been explored in detail.

**Methods:** 31 individuals with 3q29 deletion syndrome (3q29del, 61.3% male) were evaluated using two gold-standard clinical ASD evaluations: the Autism Diagnostic Observation Schedule, Second Edition (ADOS-2) and the Autism Diagnostic Interview, Revised (ADI-R). Four matched comparators for each subject were ascertained from the National Database for Autism Research. Item-level scores on the ADOS-2 and ADI-R were compared between subjects with 3q29del and matched comparators.

**Results:** Subjects with 3q29del and no ASD (3q29del-ASD) had greater evidence of social disability compared to typically developing (TD) comparison subjects across the ADOS-2. Subjects with 3q29del and ASD (3q29del+ASD) were largely indistinguishable from non-syndromic ASD (nsASD) subjects on the ADOS-2. 3q29del+ASD performed significantly better on social communication on the ADI-R than nsASD (3q29+ASD mean = 11.36; nsASD mean = 15.70; p = 0.01), and this was driven by reduced deficits in nonverbal communication (3q29+ASD mean = 1.73; nsASD mean = 3.63; p = 0.03). 3q29del+ASD reported significantly later age at first two-word phrase compared to nsASD (3q29del+ASD mean = 43.89 months; nsASD mean = 37.86 months; p = 0.01). However, speech delay was not related to the improved nonverbal communication in 3q29del+ASD.

**Limitations:** There were not enough TD comparators with ADI-R data in NDAR to include in the present analysis. Additionally, our relatively small sample size made it difficult to assess race and ethnicity effects.

**Conclusions:** 3q29del is associated with significant social disability, irrespective of ASD diagnosis. 3q29del+ASD have similar levels of social disability to nsASD, while 3q29del-ASD have significantly increased social disability compared to TD individuals. However, social communication is reasonably well-preserved in 3q29del+ASD relative to nsASD. It is critical that verbal ability and social disability be examined separately in this population to ensure equal access to ASD and social skills evaluations and services.

## Background

The 3q29 deletion is a rare (∼1 in 30,000) (1, 2) 1.6 Mb typically *de novo* deletion on chromosome 3 (hg19, chr3:195725000–197350000) (3-5). The 3q29 deletion has well-established links to multiple neurodevelopmental and neuropsychiatric disorders, including a 19-fold increased risk for autism spectrum disorder (ASD) (6-8) and a > 40-fold increased risk for schizophrenia (SZ) (9-13). Individuals with 3q29 deletion syndrome (3q29del) also may have mild to moderate intellectual disability (ID), global developmental delay (GDD), and delayed speech (3-5, 14-17). Recent efforts have led to an improved understanding of 3q29del, but nuances of some phenotypes, including social disability, require further exploration.

Previous work by our team showed that individuals with 3q29del have increased social disability relative to typically developing (TD) controls, independent of whether they meet diagnostic criteria for ASD (6). Parents of individuals with 3q29del reported significantly greater impairment than controls on several parent-report questionnaires that assessed different domains of social behavior, including social responsiveness and social communication (6). This held true even in the absence of an ASD diagnosis in the proband. However, the core measure used in this study, the Social Responsiveness Scale (SRS), can show elevated T-scores in the presence of multiple neurodevelopmental and neuropsychiatric disorders, and may not be specific to ASD (18-20). Therefore, it is unclear whether these increased scores are due to ASD-related symptoms specifically, or elevated social disability more generally. Further, this study relied on parent-report questionnaires, which can be biased (21). Thus, the elevated social disability observed in individuals with 3q29del without an ASD diagnosis may be due to the presence of co-occurring neurodevelopmental or psychiatric conditions or may simply be driven by parental bias in reporting. Alternatively, the elevated social disability may be due to undiagnosed ASD within the 3q29del population.

While studies utilizing parent-report questionnaires can provide broad insight into social disability phenotypes within the 3q29del population, a better understanding may be achieved via direct clinical evaluation. In the current study we used gold-standard diagnostic instruments administered by our team of expert clinicians to explore nuances of social disability specifically relevant to ASD within our 3q29del study participants. Specifically, we sought to compare the social disability profile from individuals with 3q29del and a diagnosis of ASD to individuals with non-syndromic ASD, as assessed by the Autism Diagnostic Observation Schedule, Second Edition (ADOS-2) and the Autism Diagnostic Interview, Revised (ADI-R). We also compared the ADOS-2 performance from individuals with 3q29del who did not qualify for an ASD diagnosis to a set of individuals without 3q29del who do not have an ASD diagnosis. Developing a better understanding of the ASD phenotype in all individuals with 3q29del will help improve clinical care for affected individuals. Further, our use of gold-standard evaluations will facilitate cross-disorder comparison to define core ASD phenotypes across genetic syndromes, as well as areas of phenotypic divergence ripe for future investigation.

## Methods

### Sample

32 individuals with 3q29del were recruited through the online 3q29 deletion registry (3q29deletion.org) (5) for in-person evaluation at the Marcus Autism Center in Atlanta, GA (22). One individual was evaluated using the ADOS-2 Module 1 and was excluded from the current analysis due to low item overlap with ADOS-2 Modules 2-4. 31 individuals with 3q29del (61.3% male) were included in the present study, ranging in age from 4.9-39.1 years (mean = 14.59 ± 8.38 years). Comparison samples of study subjects were ascertained from the National Database for Autism Research (NDAR), matched on age, sex, and ASD diagnosis status, and on race and ethnicity when possible (see Supplement for details). To maximize power, four comparison participants were selected for each 3q29del participant. The comparison sample for participants with 3q29del and a clinical ASD diagnosis (3q29del+ASD, n = 12) were individuals with a diagnosis of non-syndromic ASD (nsASD, n = 48); comparators for participants with 3q29del without a clinical ASD diagnosis (3q29del-ASD, n=19) were typically developing (TD, n = 76) individuals. Individuals in NDAR with any reported neurodevelopmental concern or diagnosis were excluded from the TD group. A description of the study sample can be found in Table 1. This study was approved by Emory University’s Institutional Review Board (IRB00064133) and Rutgers University’s Institutional Review Board (PRO2021001360).

**Table 1.** Participant demographics.

|  |  | ADOS-2 (N = 155) |  |  | ADI-R (N = 55) |  |  |
| --- | --- | --- | --- | --- | --- | --- | --- |
|  |  | 3q29del (n = 31) | Comparator (n = 124) | P value | 3q29del (n = 11) | Comparator (n = 44) | P value |
| Sex | Male n (%) | 19 (61.30) | 19 (61.30) | 1.00 | 9 (81.82) | 36 (81.82) | 1.00 |
| Age | Mean ± SD | 14.59 ± 8.38 | 14.45 ± 8.32 | 0.94 | 14.75 ± 5.11 | 14.74 ± 5.00 | 1.00 |
| Race | White n (%) | 28 (90.32) | 104 (83.87) | 1.00 | 9 (81.82) | 36 (81.82) | 1.00 |
|  | Black n (%) | 0 (0.00) | 1 (0.81) |  | 0 (0.00) | 0 (0.00) |  |
|  | Other n (%) | 3 (9.68) | 16 (12.90) |  | 2 (18.18) | 7 (15.91) |  |
|  | Unknown/not reported n (%) | 0 (0.00) | 3 (2.42) |  | 0 (0.00) | 1 (2.27) |  |
| Ethnicity | Hispanic/Latino n (%) | 1 (3.23) | 3 (2.42) | 1.88E-06 | 0 (0.00) | 0 (0.00) | 0.18 |
|  | Not Hispanic/Latino n (%) | 30 (96.77) | 71 (57.26) |  | 11 (100.00) | 34 (77.27) |  |
|  | Unknown/not reported n (%) | 0 (0.00) | 50 (40.32) |  | 0 (0.00) | 10 (22.73) |  |
| Diagnosis | ASD n (%) | 12 (38.71) | 48 (38.71) | 1.00 | 11 (100.00) | 44 (100.00) | 1.00 |

Demographic data collected through the online 3q29 registry (for 3q29del participants) or reported to NDAR (for comparator participants). P values were calculated with Fisher's exact test (diagnosis, sex, race, and ethnicity) and two-sample t-test (age). For race, "Other" includes Hawaiian or Pacific Islander (n = 1 comparator ADOS-2), more than one race (n = 3 3q29del ADOS-2, 11 comparator ADOS-2, 2 3q29del ADI-R, 7 comparator ADI-R), American Indian or Alaska Native (n = 2 comparator ADOS-2), and Asian (n = 2 comparator ADOS-2)

### Measures

Gold-standard clinical evaluation of ASD symptomatology was performed using the Autism Diagnostic Observation Schedule, Second Edition (23) (ADOS-2, n = 12 3q29del+ASD, 19 3q29del-ASD, 48 nsASD, 76 TD) and Autism Diagnostic Interview, Revised (24, 25) (ADI-R, n = 11 3q29del+ASD, 44 nsASD). Additional detail regarding the administration and scoring of the ADOS-2 and ADI-R can be found in the Supplemental Methods.

#### ADOS-2

The ADOS-2 is a semi-structured, play-based, observational assessment of social interaction, communication, play and imagination skills, and repetitive behaviors. There are five different modules for the ADOS-2 corresponding to the subject’s age and language level: the Toddler Module is used for nonverbal or minimally verbal toddlers between 12 and 30 months, Module 1 is used for nonverbal or minimally verbal individuals who are at least 31 months of age, Module 2 for individuals who speak in words and phrases of any age, Module 3 for individuals who are verbally fluent up to older adolescence, and Module 4 for individuals who are verbally fluent and are older adolescents or adults. Within each module, items are grouped into four categories: language and communication, reciprocal social interaction, play/imagination, and stereotyped behaviors and restricted interests. Higher domain and item scores correspond to greater impairment.

#### ADI-R

The ADI-R is a semi-structured interview between a parent and clinician focused on the early developmental history and current and lifetime behavior of the subject. The diagnostic algorithm of the ADI-R was used in the present study, which focuses on symptom presentation in early childhood. Items on the ADI-R are grouped into four domains for scoring: qualitative abnormalities in reciprocal social interaction; qualitative abnormalities in communication; restricted, repetitive, and stereotyped patterns of behavior; and abnormality of development evident at or before 36 months. The first three domains are further divided into sub-domains that capture different aspects of the domain. Higher domain, sub-domain, and item scores indicate greater symptom severity.

Despite comprehensive searches in NDAR, there were not enough TD subjects with ADI-R data, so a comparison of ADI-R performance between 3q29del cases without ASD and TD subjects was not performed.

#### Analysis

All analyses were performed in R version 4.0.4 (26). For the ADOS-2, analyses were performed at the domain- and item-level. Domain-level Social Affect (SA) and Restricted and Repetitive Behaviors (RRB) raw scores were calculated according to the module-appropriate algorithms and were converted to Calibrated Severity Scores (CSS) for analysis across modules (23, 27, 28). ADOS-2 item-level analyses were performed on a harmonized set of items with consistent names, descriptions, and scoring guidelines across ADOS-2 Modules 2-4 (Table 2). Exceptions for item matching were made if wording differences were due to the developmental nature of the module, but the item was capturing the same ability. Items from the “Other Abnormal Behaviors” section were excluded, as these items do not assess ASD-specific behaviors. Item-level harmonization (Table 2) was confirmed by trained clinicians, one of whom is a certified ADOS-2 Trainer (CK). For the ADI-R, domains were calculated according to the companion scoring algorithm (24, 25). Sub-domain and item-level analyses were performed on any domains that showed a significant difference between 3q29del+ASD and nsASD participants.

**Table 2.** ADOS-2 item harmonization.

| ADOS-2 Sections | Core Item Number | Item Name |  |  | % Completed |
| --- | --- | --- | --- | --- | --- |
|  |  | Module 2 | Module 3 | Module 4 |  |
| Language and Communication |  | Overall level of non-echoed spoken language | Overall level of non-echoed spoken language | Overall level of non-echoed spoken language |  |
|  | 1 | **Speech abnormalities associated with autism (intonation/volume/rhythm /rate)** | **Speech abnormalities associated with autism (intonation/volume/rhythm /rate)** | **Speech abnormalities associated with autism (intonation/volume/rhythm /rate)** | **92.90%** |
|  | 2 | **Immediate echolalia** | **Immediate echolalia** | **Immediate echolalia** | **98.06%** |
|  | 3 | **Stereotyped/idiosyncratic use of words or phrases** | **Stereotyped/idiosyncratic use of words or phrases** | **Stereotyped/idiosyncratic use of words or phrases** | **98.71%** |
|  | 4 | **Conversation** | **Conversation** | **Conversation** | **95.48%** |
|  |  | Pointing | Asks for information | Asks for information |  |
|  | 5 | **Descriptive, conventional, instrumental, or informational gestures** | **Descriptive, conventional, instrumental, or informational gestures** | **Descriptive, conventional, instrumental, or informational gestures** | **98.71%** |
|  |  |  | Offers information | Offers information |  |
|  |  |  | Reporting of events | Reporting of events |  |
|  |  |  |  | Emphatic or emotional gestures |  |
| Reciprocal Social Interaction | 6 | **Unusual eye contact** | **Unusual eye contact** | **Unusual eye contact** | **98.06%** |
|  | 7 | **Facial expressions directed to others** | **Facial expressions directed to examiner** | **Facial expressions directed to examiner** | **94.84%** |
|  | 8 | **Shared enjoyment in interaction** | **Shared enjoyment in interaction** | **Shared enjoyment in interaction** | **97.42%** |
|  |  | Response to name | Language production and linked nonverbal | Language production and linked nonverbal |  |
|  |  |  | communication | communication |  |
|  |  | Showing | Insight into typical social situations and relationships | Insight into typical social situations and relationships |  |
|  |  | Spontaneous imitation of joint attention | Comments on others' emotions/empathy | Comments on others' emotions/empathy |  |
|  | 9 | Quality of social overtures | Quality of social overtures | Quality of social overtures | 98.06% |
|  | 10 | Amount of social overtures/maintenance of attention (examiner) | Amount of social overtures/maintenance of attention | Amount of social overtures/maintenance of attention | 83.87% |
|  |  | Amount of social overtures/maintenance of attention (parent/caregiver) |  | Communication of own affect |  |
|  | 11 | Quality of social response | Quality of social response | Quality of social response | 98.06% |
|  | 12 | Amount of reciprocal social communication | Amount of reciprocal social communication | Amount of reciprocal social communication | 98.06% |
|  | 13 | Overall quality of rapport | Overall quality of rapport | Overall quality of rapport | 95.48% |
|  |  | Response to joint attention |  | Responsibility |  |
| Play/Imagination | 14 | Imagination/creativity | Imagination/creativity | Imagination/creativity | 98.06% |
|  |  | Functional play with objects |  |  |  |
| Stereotyped Behaviors and Restricted Interests | 15 | Unusual sensory interest in play material/person | Unusual sensory interest in play material/person | Unusual sensory interest in play material/person | 98.71% |
|  | 16 | Hand and finger and other complex mannerisms | Hand and finger and other complex mannerisms | Hand and finger and other complex mannerisms | 98.71% |
|  | 17 | Self-injurious behavior | Self-injurious behavior | Self-injurious behavior | 98.06% |
|  | 18 | Unusually repetitive interests or stereotyped behaviors | Excessive interest in or references to unusual or highly specific topics or objects or repetitive behaviors | Excessive interest in or references to unusual or highly specific topics or objects or repetitive behaviors | 95.48% |
|  |  |  | Compulsions or rituals | Compulsions or rituals |  |

ADOS-2 Modules 2-4 item harmonization. Harmonized items are bolded. The percentage of study subjects that were scored on each harmonized item is noted.

Statistical analysis of demographic variables was performed using Fisher’s exact test and two sample *t* test implemented using the stats R package (26). To compare ADOS-2 and ADI-R variables between 3q29del and comparator groups, generalized linear models, cumulative link models, and simple linear models were implemented using the stats (26) and ordinal (29) R packages. For ADOS-2 items without sufficient variation, Wilcoxon rank sum tests were implemented using the stats R package (26). Data visualization was performed using the plotly R package (30).

## Results

### ADOS-2 reveals nuances of social disability in individuals with 3q29del and ASD

The ADOS-2 revealed no statistically significant differences between 3q29del participants with ASD and nsASD comparators on the SA (3q29del+ASD mean = 8.25 ± 0.87; nsASD mean = 7.26 ± 1.84; p > 0.05) or RRB (3q29del+ASD mean = 7.25 ± 2.26; nsASD mean = 6.89 ± 2.81; p > 0.05) domains (Figure 1A-B). When comparing specific ADOS-2 items, 3q29del participants with ASD scored significantly higher (worse) than nsASD participants on items related to reciprocal social interaction: unusual eye contact (item 6, 3q29del+ASD mean = 2.00 ± 0.00; nsASD mean = 1.40 ± 0.92; p = 0.03), facial expressions directed towards others or the examiner (item 7, 3q29del+ASD mean = 1.33 ± 0.49; nsASD mean = 1.00 ± 0.43; p = 0.02), and amount of social overtures and maintenance of attention during the assessment (item 10, 3q29del+ASD mean = 1.75 ± 0.87; nsASD mean = 1.18 ± 0.91; p = 0.04) (Figure 1C). These data suggest that reciprocal social interaction may be a specific area of severe vulnerability for individuals with 3q29del and ASD, and may require targeted intervention.

**Figure 1.**
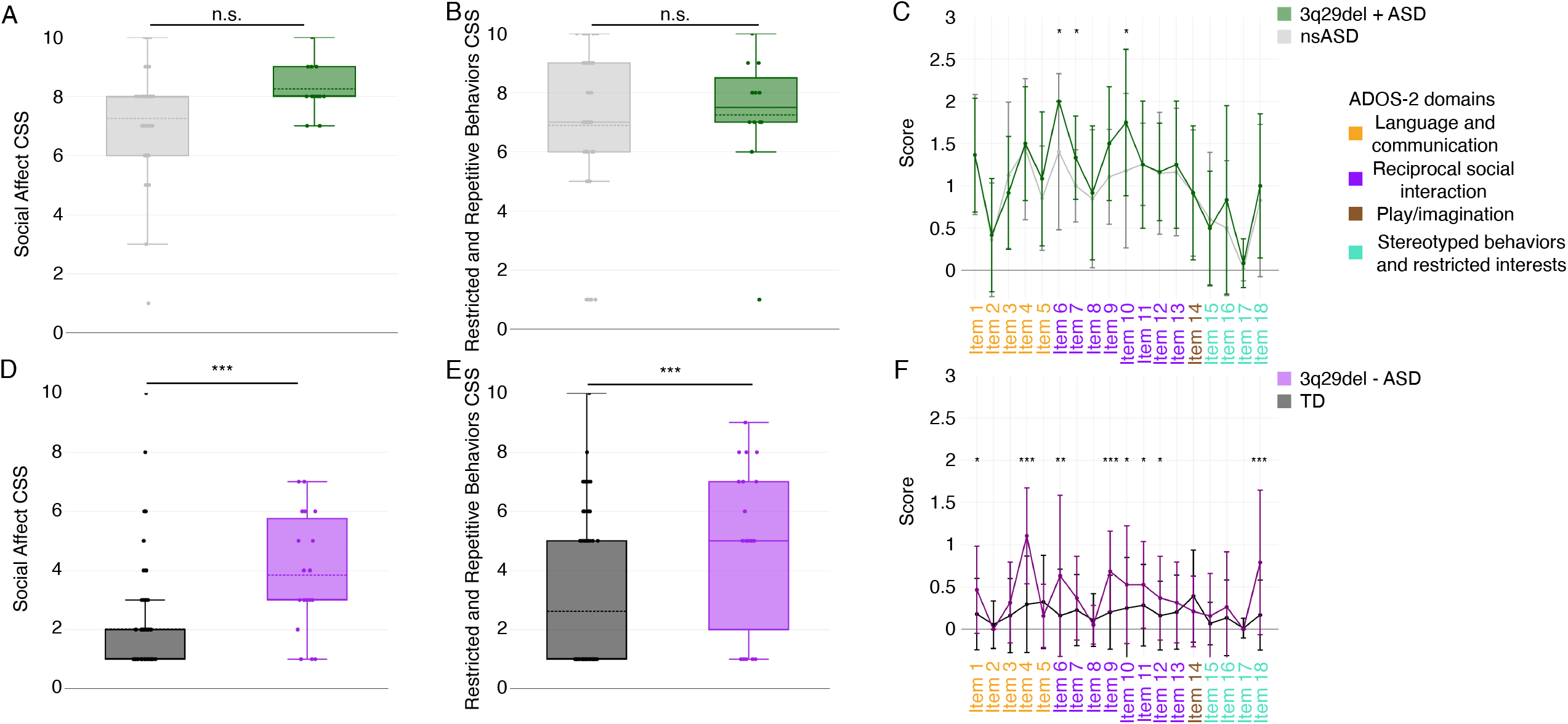
ADOS-2 domain and item scores. A-B) Social affect (SA, A) and restricted and repetitive behaviors (RRB, B) calibrated severity scores (CSS) for individuals with 3q29del and ASD (n = 12) and nsASD comparators (n = 48), showing no significant difference in scores. C) ADOS-2 harmonized item scores for individuals with 3q29del and ASD (n = 12) and nsASD comparators (n = 48), showing significantly increased scores in individuals with 3q29del and ASD on items 6, 7, and 10. D-E) SA (D) and RRB (E) CSS for individuals with 3q29del and no ASD (n = 19) and TD comparators (n = 76), showing significantly increased scores in individuals with 3q29del and no ASD. F) ADOS-2 harmonized item scores for individuals with 3q29del and no ASD (N = 19) and TD comparators (n = 76), showing significantly increased scores in individuals with 3q29del and no ASD on items 1, 4, 6, 9, 10, 11, 12, and 18. n.s., p > 0.05; *, p < 0.05; ***, p < 0.001

### ADOS-2 reveals increased social disability in individuals with 3q29del and no ASD

Within the non-ASD group, 3q29del study subjects without an ASD diagnosis scored significantly higher than TD comparators on the SA (3q29del-ASD mean = 3.84 ± 1.95; TD mean = 2.01 ± 1.77; p = 4.24E-6) and RRB (3q29del-ASD mean = 5.00 ± 2.75; TD mean = 2.62 ± 2.46; p = 0.0008) domains (Figure 1D-E). At the item-level of analysis, 3q29del participants without ASD scored significantly higher than TD participants on 8 items: speech abnormalities associated with ASD (item 1, 3q29del-ASD mean = 0.47 ± 0.52; TD mean = 0.18 ± 0.42; p = 0.02), conversation (item 4, 3q29del-ASD mean = 1.11 ± 0.57; TD mean = 0.30 ± 0.57; p = 4.22E-7), unusual eye contact (item 6, 3q29del-ASD mean = 0.63 ± 0.96; TD mean = 0.16 ± 0.55; p = 0.007), the quality of social overtures (item 9, 3q29del-ASD mean = 0.68 ± 0.48; TD mean = 0.20 ± 0.44; p = 0.0001), the amount of social overtures and maintenance of attention (item 10, 3q29del-ASD mean = 0.53 ± 0.70; TD mean = 0.25 ± 0.60; p = 0.03), the quality of social response (item 11, 3q29del-ASD mean = 0.53 ± 0.51; TD mean = 0.28 ± 0.48; p = 0.04), the amount of reciprocal social communication (item 12, 3q29del-ASD mean = 0.37 ± 0.50; TD mean = 0.16 ± 0.41; p = 0.04), and repetitive interests and repetitive or stereotyped behaviors evident during the evaluation (item 18, 3q29del-ASD mean = 0.79 ± 0.85; TD mean = 0.17 ± 0.41; p = 0.0003) (Figure 1F). These data indicate that for individuals with 3q29del, significant social disability may be present even in the absence of an ASD diagnosis, and social skills interventions are warranted.

### ADI-R shows 3q29del participants with ASD perform better on social communication than nsASD

To further define nuances of the social disability phenotype within our population of individuals with 3q29del and ASD, we examined data from the ADI-R. Of the four core domains assessed by the ADI-R, only one domain was significantly different between 3q29del participants with ASD and nsASD participants (Figure 2). 3q29del participants with ASD scored significantly lower (better) than nsASD participants on domain B, corresponding to “qualitative abnormalities in communication” (3q29+ASD mean = 11.36 ± 5.63; nsASD mean = 15.70 ± 4.75; p = 0.01; Figure 2B), indicating less impairment in social communication in individuals with 3q29del and ASD compared to nsASD subjects.

**Figure 2.**
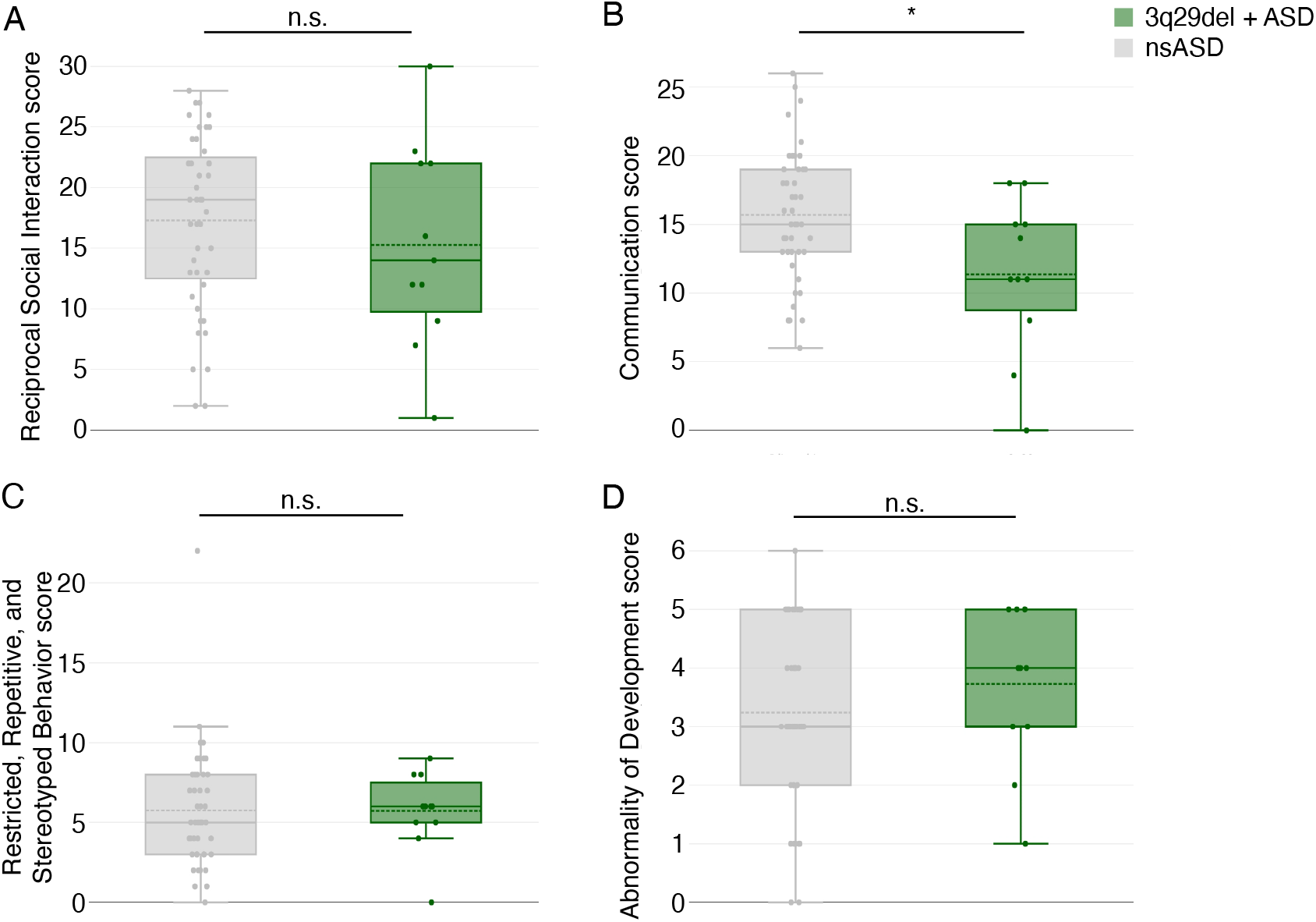
ADI-R domain scores. A) ADI-R domain A scores, showing no significant difference between individuals with 3q29del and ASD (n = 11) and nsASD comparators (n = 44). B) ADI-R domain B scores, showing significantly lower scores in individuals with 3q29del and ASD (n = 11) compared to nsASD comparators (n = 44). C) ADI-R domain C scores, showing no significant difference between individuals with 3q29del and ASD (n = 11) and nsASD comparators (n = 44). D) ADI-R domain D scores, showing no significant difference between individuals with 3q29del and ASD (n = 11) and nsASD comparators (n = 44). n.s., p > 0.05; *, p < 0.05

At the domain level, it was unclear whether the significantly reduced domain B score in individuals with 3q29del and ASD was due to decreased scores across multiple factors, or a specific difference in a single sub-domain. Analysis of the four sub-domains that comprise domain B revealed that only one was significantly lower in participants with 3q29del and ASD compared to nsASD (Figure 3A-D). 3q29del participants with ASD scored significantly lower than nsASD participants on sub-domain B1, “lack of, or delay in, spoken language and failure to compensate through gesture” (3q29+ASD mean = 1.73 ± 2.45; nsASD mean = 3.63 ± 2.54; p = 0.03; Figure 3A). Sub-domain B1 is comprised of four items: “pointing to express interest” (item 42), “nodding” (item 43), “head shaking” (item 44), and “conventional/instrumental gestures” (item 45). Within sub-domain B1, participants with 3q29del and ASD scored slightly lower than nsASD participants on all four items, but the differences were not statistically significant (Figure 3E).

**Figure 3.**
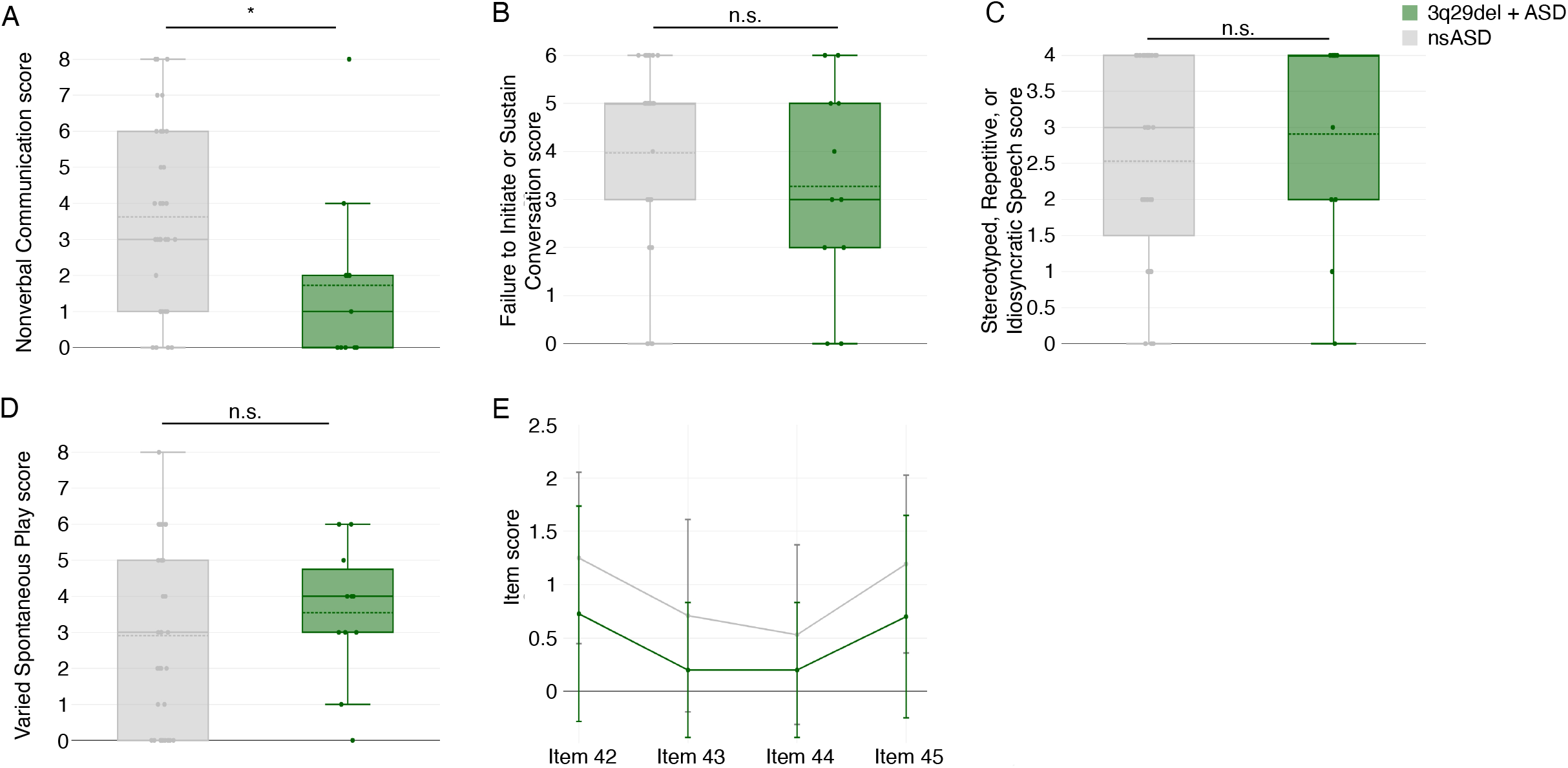
ADI-R sub-domain and item scores. A) ADI-R sub-domain B1 scores, showing significantly lower scores in individuals with 3q29del and ASD (n = 11) compared to nsASD comparators (n = 32). B) ADI-R sub-domain B2 scores, showing no significant difference between individuals with 3q29del and ASD (n = 11) and nsASD comparators (n = 32). C) ADI-R sub-domain B3 scores, showing no significant difference between individuals with 3q29del and ASD (n = 11) and nsASD comparators (n = 32). D) ADI-R sub-domain B4 scores, showing no significant difference between individuals with 3q29del and ASD (n = 11) and nsASD comparators (n = 32). E) ADI-R sub-domain B1 item scores for individuals with 3q29del and ASD (n = 11) and nsASD comparators (n = 32), showing no significant difference on any sub-domain items. n.s., p > 0.05; *, p < 0.05

### Speech delay is not related to higher nonverbal communication scores in 3q29del

We hypothesized that the use of nonverbal communication strategies may be related to the degree of speech delay in our study participants. Specifically, we asked whether individuals with a more significant speech delay compensated for their lack of speech through the use of stronger nonverbal communication skills. To investigate this hypothesis, we analyzed the age of first single word and the age of first two-word phrases as recorded on the ADI-R, in relation to ADI-R domain B (social communication) and sub-domain B1 (nonverbal communication) scores. We found that there was no genotype difference in age at first single word (3q29del+ASD mean = 21.45 ± 13.48 months; nsASD mean = 20.93 ± 12.75 months; p = 0.88; Figure 4A). After adjusting for genotype, age at first single word was significantly associated with the nonverbal communication score (p = 0.03; Figure 4B, Table S3). In genotype-specific analyses, age at first single word was significantly associated with nonverbal communication scores in nsASD comparators (p = 0.005; Figure 4C, Table S3), but the association was not statistically significant in individuals with 3q29del and ASD (Figure 4D, Table S3). On average, individuals with 3q29del and ASD spoke their first two-word phrase approximately 6 months later than nsASD comparators (3q29del+ASD mean = 43.89 ± 31.64 months; nsASD mean = 37.86 ± 20.76 months; p = 0.01; Figure 4E). However, age at first two-word phrase was not significantly associated with social communication or nonverbal communication scores (Table S3). Taken together, these data demonstrate that speech delay cannot account for the less-impacted nonverbal communication in our 3q29del participants.

**Figure 4.**
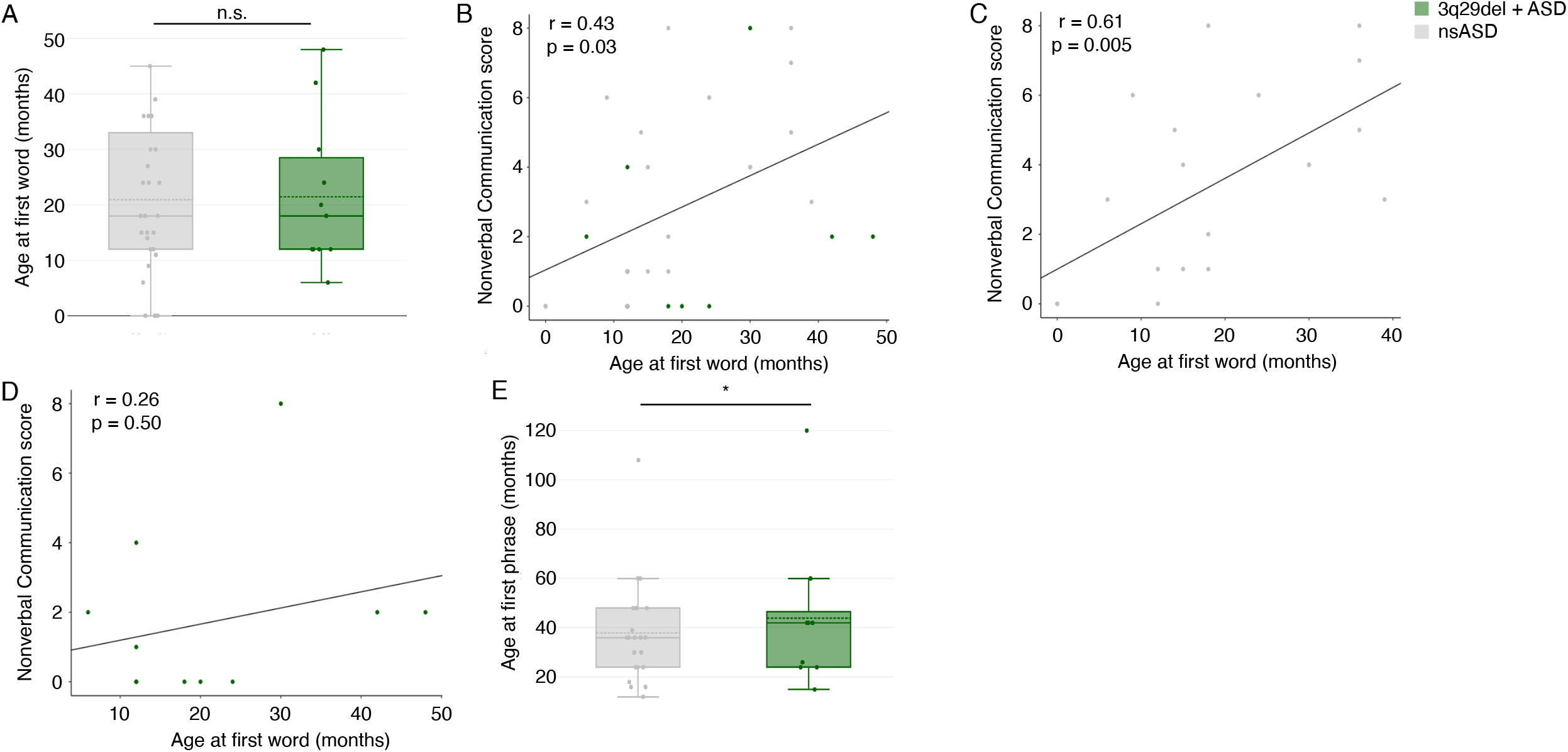
Speech delay and ADI-R nonverbal communication scores. A) Age at first single word (months) is not significantly different between individuals with 3q29del and ASD (n = 11) and nsASD comparators (n = 32). B) Relationship between age at first single word (months) and sub-domain B1 scores in the full dataset (3q29del+ASD n = 11; nsASD n = 32), showing a significant positive relationship. C) Relationship between age at first single word (months) and sub-domain B1 scores in nsASD comparators (n = 32), showing a significant positive relationship. D) Relationship between age at first single word (months) and sub-domain B1 scores in individuals with 3q29del and ASD (n = 11), showing no significant relationship. E) Age at first two-word phrase (months) is significantly later in individuals with 3q29del and ASD (n = 11) compared to nsASD comparators (n = 32). n.s., p > 0.05; *, p < 0.05

## Discussion

The present study is the first to use gold-standard ASD evaluations to assess features of social disability in individuals with 3q29del. Individuals with 3q29del and ASD are largely indistinguishable from individuals with nsASD on both the ADOS-2 (Figure 1A-C) and ADI-R (Figures 2 and 3), with some minor differences on specific items. In contrast, individuals with 3q29del and no ASD had substantially more social disability than TD comparators on the ADOS-2, with scores for the SA and RRB domains as well as significantly elevated scores on nearly 50% of the 18 harmonized ADOS-2 items (Figure 1D-F). These findings are consistent with previous work by our group, in which we found significant social disability within our 3q29del patient population independent of ASD diagnosis relative to TD controls (6).

There were some specific areas of divergence between individuals with 3q29del and ASD and nsASD comparators. On the ADOS-2, individuals with 3q29del and ASD scored significantly higher than nsASD comparators on 3 harmonized items related to social reciprocity during the evaluation; namely, unusual eye contact, facial expressions directed toward the examiner, and the frequency of social overtures and maintenance of attention during the assessment. These data suggest that individuals with 3q29del and ASD are uniquely impaired in these areas, with greater impairment than is observed in children with nsASD. On the ADI-R, individuals with 3q29del and ASD scored significantly lower than nsASD comparators on the social communication domain, specifically on nonverbal communication. This indicates that social communication, particularly nonverbal communication, is relatively well-preserved in individuals with 3q29del and ASD relative to individuals with nsASD. A previous analysis of the Social Communication Questionnaire (SCQ) by our team revealed that our 3q29del study population as a whole had mean scores in the normal range, and individuals with 3q29del and ASD had scores only slightly above the clinical cutoff (6). Additionally, clinical evaluations by our team found that verbal IQ in 3q29del is significantly higher than nonverbal IQ (16). Together, these data support our current finding of social communication as a relative strength within our sample of individuals with 3q29del and ASD, while social reciprocity is an area of vulnerability and a potential target for intervention.

The high prevalence of social disability in individuals with 3q29del and no ASD diagnosis is a critical consideration in this population. Typically, a diagnosis of ASD is required to qualify for social skills interventions and other therapeutic services. However, a majority of individuals with 3q29del do not qualify for an ASD diagnosis, even in the presence of substantial social disability. Previous work by our team (6) in combination with the present work highlights the critical need for early, gold-standard ASD evaluation for individuals diagnosed with 3q29del. Further, the high degree of social disability independent of ASD diagnosis demonstrates that a diagnosis of 3q29del alone should be sufficient for an individual to qualify for social skills interventions and other early intervention strategies. The high prevalence of social disability associated with 3q29del also suggests that the 3q29 deletion impacts key biological pathways relevant to social function. Thus, the 3q29 deletion may serve as a window to understanding the molecular pathology underlying social disability.

Previous work by our group using parent-report questionnaires identified a high burden of social disability within our 3q29del study population; however, average scores for social communication were within the normal range (6). The present study supports this finding and further suggests that social communication is better preserved in individuals with 3q29del and ASD than other aspects of social behavior, and nonverbal communication ability appears to drive this preservation. We hypothesized that early speech delay in our 3q29del participants may incentivize the development of nonverbal communication skills as a compensatory mechanism. Study subjects with 3q29del and ASD had significantly later age at first two-word phrase compared to nsASD comparators (Figure 4E); however, this delay was not associated with measures of nonverbal communication in our 3q29del participants. Based on these data, speech delay alone is not sufficient to explain the relatively preserved nonverbal communication in individuals with 3q29del and ASD compared to nsASD comparators. Further, because nonverbal social communication is relatively well-preserved in individuals with 3q29del, other ASD symptoms may be overlooked. This highlights the critical need for gold-standard ASD evaluations as standard of care for all individuals diagnosed with 3q29del.

Taken together, the data presented in the current study reveal substantial social disability within the 3q29del population as assessed by gold-standard ASD evaluations. This finding is supported by a previous study by our team that examined parent-report questionnaires that focused on social disability phenotypes (6). Coupled with our previous work, the present findings emphasize the need for early gold-standard ASD evaluation for all individuals with a diagnosis of 3q29 deletion syndrome. Additionally, these data support the need for social skills interventions within the 3q29del population, even in the absence of a clinical ASD diagnosis. Individuals with 3q29del and no ASD diagnosis may be uniquely vulnerable to social disability because individuals without an ASD diagnosis are traditionally not prioritized for social skills interventions. By prioritizing early social skills therapies for all individuals with 3q29del, we may improve the social function of the entire population of individuals with 3q29del, rather than only focusing on those that qualify for a clinical ASD diagnosis.

### Limitations

While this study contributes valuable information to our understanding of social disability within the 3q29del population, it is not without limitations. There were some challenges with identifying comparators in NDAR. First, we were unable to identify a sufficient number of TD comparators with an ADI-R within the database, which caused us to restrict our ADI-R analysis to only individuals with 3q29del and ASD and nsASD comparators. Second, the TD comparators that were identified for the ADOS-2 analyses may not truly be TD. While no ASD or other clinical diagnosis was noted, it is unclear whether some of these individuals received an ADOS-2 evaluation because of other developmental concerns that did not meet the threshold for diagnosis. However, even if there is some developmental concern within the TD group, individuals with 3q29del and no ASD diagnosis still showed significantly more impairment on the ADOS-2. This suggests that this limitation may not have substantially impacted the core findings of the present study. Finally, we were unable to assess race and ethnicity effects in the present study, due to small sample size and a lack of diversity within the 3q29del registry. Future effort is required to reach a more diverse population of individuals with 3q29del. Additionally, systemic changes are required to address long-standing disparities in the access to and utilization of genetic services, which currently may contribute to under-diagnosis of genetic and genomic syndromes like 3q29del within minority populations (31-35).

## Conclusions

The present study is the first to examine nuances of social disability within the 3q29del population using gold-standard ASD evaluations. We also present a harmonized approach for analyzing item-level data across ADOS-2 modules 2, 3, and 4. We find significant social disability present in our 3q29del study population; individuals with 3q29del and ASD have a similar degree of social disability to nsASD comparators, while individuals with 3q29del and no ASD have substantially greater social disability than TD comparators. Additionally, individuals with 3q29del and ASD have relatively well-preserved social communication, specifically nonverbal communication, and relatively impaired social reciprocity, compared to nsASD comparators. We hypothesize that ASD and social disability may be under-appreciated in some cases of 3q29del due to the relatively preserved verbal ability and social communication within this population. This hypothesis is supported by anecdotal reports from parents of children with 3q29del, who recount having their concerns about ASD in their child dismissed because their child was considered “too verbal” by their clinician. These reports indicate that improved clinician education surrounding the distinction between verbal ability and social disability may be merited to ensure equal access to ASD evaluations and social skills interventions for individuals with 3q29del. Based on these data and previous work by our team (6, 17), we recommend that all individuals with 3q29del should receive gold-standard ASD evaluations as standard of care, regardless of the individual’s verbal ability. Early diagnosis of social deficits and early therapeutic intervention within this patient population will be the most effective way to improve future outcomes and quality of life for individuals with 3q29del and their families.

## Data Availability

The datasets used and/or analyzed during the current study are available from the corresponding author on reasonable request.

## List of abbreviations

3q29del: 3q29 deletion syndrome
ASD: autism spectrum disorder
SZ: schizophrenia
ID: intellectual disability
GDD: global developmental delay
TD: typically developing
SRS: Social Responsiveness Scale
ADOS-2: Autism Diagnostic Observation Schedule, Second Edition
ADI-R: Autism Diagnostic Interview, Revised
NDAR: National Database for Autism Research
nsASD: non-syndromic autism spectrum disorder
SA: social affect
RRB: restricted and repetitive behaviors
CSS: calibrated severity score
SCQ: Social Communication Questionnaire

## Declarations

### Ethics approval and consent to participate

This study was approved by Emory University’s Institutional Review Board (IRB00064133) and Rutgers University’s Institutional Review Board (PRO2021001360). All study subjects gave informed consent prior to participating in this study.

### Consent for publication

Not applicable.

### Authors’ contributions

RMP identified the ADI-R comparator dataset in NDAR; performed the statistical analysis; produced all figures; produced tables 1, S2, and S3; and wrote the manuscript. JEP identified the ADOS-2 comparator dataset in NDAR, produced tables 2 and S1, performed ADOS-2 item harmonization, and calculated ADOS-2 domain scores. CK and CAS helped with ADOS-2 item harmonization. CK, CAS, and SPW performed the ADOS-2 and ADI-R evaluations of 3q29 deletion syndrome study participants. TLB, JFC, CK, MMM, CAS, EFW, and SPW helped with data interpretation. JGM edited the manuscript and provided guidance on analyzing and interpreting data. JGM was the principal investigator responsible for study direction. All authors participated in commenting on the drafts and have read and approved the final manuscript.

## Acknowledgements

We gratefully acknowledge our study population, the 3q29 deletion community, for their participation and commitment to research.

