## Supplemental Information for "The clinical phenotype of autism spectrum disorder in individuals with 3q29 deletion syndrome"

**Supplemental Methods**

*Matching comparators for 3q29del participants*

Four comparators for each 3q29del participant were ascertained from the National Database for Autism Research (NDAR, Table S1). Comparators for participants with 3q29del and a clinical ASD diagnosis (3q29del+ASD, n = 12) were individuals with non-syndromic ASD (nsASD); comparators for participants with 3q29del without a clinical ASD diagnosis (3q29del-ASD, n=19) were TD individuals. All comparators were matched on age, sex, and ASD diagnosis status, and were matched on race and ethnicity when possible. For matching on age, individuals under 18 were matched on exact age, meaning a match for an 8 year old individual would be between 8 years, 0 months, and 8 years, 11 months. Individuals over 18 were matched within a two year range, centered on their age, meaning a match for a 24 year old individual would be between 23 years, 0 months, and 25 years, 11 months. Exceptions for age matching were made for 7 participants in the ADOS-2 data set (Table S2). No exceptions for age matching were made in the ADI-R data set.

When possible, the same comparators were used in the ADOS-2 and ADI-R datasets. However, there was limited overlap between the comparator data sets due to differences in data availability for studies in NDAR (Figure S1).

*ADOS-2*

The ADOS-2 is a semi-structured, standardized, observational assessment of social interaction, communication, play and imagination skills, and repetitive behaviors. During the assessment, the subject is placed in different play-based situations. The clinician does not provide the subject with any instructions or guidance in order to observe the subject’s natural behaviors. There are five different modules for the ADOS-2 corresponding to the subject’s age and language level: the Toddler Module is used for nonverbal or minimally verbal toddlers between 12 and 30 months, Module 1 is used for nonverbal or minimally verbal individuals who are at least 31 months of age, Module 2 for individuals who speak in words and phrases of any age, Module 3 for individuals who are verbally fluent up to older adolescence, and Module 4 for individuals who are verbally fluent and are older adolescents or adults. Within each module, items are grouped into four categories: language and communication, reciprocal social interaction, play/imagination, and stereotyped behaviors and restricted interests. A subset of the scored items are used to calculate two domain scores, according to a standard algorithm (1). The domains are Social Affect (SA) and Restricted and Repetitive Behavior (RRB). Higher domain and item scores correspond to greater impairment.

For analysis, SA and RRB domain scores were calculated according to the companion scoring algorithms (1, 2). Because the items used to calculate the SA and RRB domain scores vary between modules, the domain scores were converted to Calibrated Severity Scores (CSS) for comparison across modules (1-3). Possible CSS values range from 0 to 10. Item-level analyses were performed on core items across modules (Table 2). Possible item scores range from 0 to 3. For item-level analysis, scores between 0 and 3 were preserved and scores of 7, 8, or 9 were dropped.

*ADI-R*

The ADI-R is a semi-structured interview between a parent and clinician focused on the early developmental history and current and lifetime behavior of the subject. The diagnostic algorithm of the ADI-R was used in the present study, which focuses on symptom presentation in early childhood. Items on the ADI-R are grouped into four domains for scoring: qualitative abnormalities in reciprocal social interaction; qualitative abnormalities in communication; restricted, repetitive, and stereotyped patterns of behavior; and abnormality of development evident at or before 36 months. The first three domains are further divided into sub-domains that capture different aspects of the domain. Scores are calculated according to the companion scoring algorithm (4, 5). Higher domain, sub-domain, and item scores indicate greater symptom severity.

For analysis, domain scores were calculated according to the companion scoring algorithm (4, 5). Analysis was performed at the domain level first; any domains showing a significant difference between individuals with 3q29del and ASD and nsASD comparators were broken into sub-domains. If any sub-domains showed a significant difference, the individual items of that sub-domain were examined. The score range for domains and sub-domains varies according to the number of items included in each domain. Item-level scores range from 0 to 3. For item-level analysis, scores between 0 and 3 were preserved and scores of 7, 8, or 9 were dropped.

**Table S1. NDAR data collections used to construct comparator datasets.**

| NDAR collection ID | NDAR collection title | ADOS-2 records (n) | ADI-R records (n) |
| --- | --- | --- | --- |
| 1 | UIC ACE: Translational Studies of Insistence on Sameness in Autism | 2 | 2 |
| 1889 | ASD Mathematical Cognition: A Cognitive and Systems Neuroscience Approach | 0 | 5 |
| 1985 | Evaluating the Time-Dependent Unfolding of Social Interactions in Children with Autism | 2 | 0 |
| 2001 | Influence of Attention and Arousal on Sensory Abnormalities in ASD | 0 | 2 |
| 2013 | Magnetoencephalographic studies of lexical processing and abstraction in autism | 1 | 0 |
| 2021 | Multimodal Developmental Neurogenetics of Females with ASD | 13 | 16 |
| 2025 | Minimally Verbal ASD: From Basic Mechanisms to Innovative Interventions | 1 | 1 |
| 2026 | Biomarkers of Developmental Trajectories and Treatment in ASD | 0 | 2 |
| 2030 | Electrophysiological Response to Executive Control Training in Autism | 1 | 4 |
| 2053 | Autism Spectrum Disorder: Birth Cohort 1976-2000, Epidemiology and Adult | 17 | 0 |
| 2066 | The CHARGE Study: Childhood Autism Risks From Genetics and the Environment | 3 | 0 |
| 2093 | Sporadic Mutations and Autism Spectrum Disorders | 4 | 0 |
| 2116 | Improving Transition Outcomes in ASD using COMPASS | 1 | 0 |
| 2120 | Adapting a Parent Advocacy Program to Improve Transition for Youth With Autism | 3 | 0 |
| 2179 | Neural markers of shared gaze during simulated social interactions in ASD \-Modal Automated Assessment of Behavior during Social Interactions in Children with ASD | 0 | 2 |
| 2266 | Inhibitory dysfunction in autism | 0 | 2 |
| 2282 | Cognitive Enhancement Therapy for Adult Autism Spectrum Disorder | 12 | 4 |
| 2285 | Integrity and Dynamic Processing Efficiency of Networks in ASD | 1 | 1 |
| 2288 | The Autism Biomarkers Consortium for Clinical Trials | 11 | 0 |
| 2292 | Molecular Mechanisms of Atypical Habituation in Autism Spectrum Disorders | 9 | 0 |
| 2293 | Components of Emotional Processing in Toddlers with ASD | 1 | 0 |
| 2368 | Clinical and Immunological Investigations of Subtypes of Autism | 13 | 0 |
| 2421 | Optimizing Prediction of Social Deficits in Autism Spectrum Disorders | 6 | 0 |
| 2441 | Structural and Functional Characteristics of XYY - Relationship to ASD | 1 | 0 |
| 2471 | A Simultaneous PET-MR Study of Striatal Dopamine Binding in Autism | 1 | 0 |
| 2706 | Thalamic activity and structure and surface neural oscillations in autism | 4 | 0 |
| 2711 | Motor abnormalities and functional brain mechanisms in autism spectrum disorder | 1 | 2 |
| 2761 | Charting the trajectory of executive control in autism in order to optimize delivery of intervention | 5 | 0 |
| 2771 | Emergent Gaze Perception in Autism Spectrum Disorder | 2 | 0 |
| 2778 | Heterogeneity in Autism Spectrum Disorders: Biological Mechanisms, Trajectories, and Treatment Response | 1 | 1 |
| 2821 | Cellular, molecular, and functional imaging approaches to understanding early neurodevelopment in autism | 5 | 0 |
| 2866 | Investigating Social Competence in Youth with Autism: A Multisite RCT | 1 | 0 |
| 2928 | Understanding Attentional Strengths and Weaknesses in Autism Spectrum Disorder | 2 | 0 |

List of NDAR data collections used to construct ADOS-2 and ADI-R comparator datasets.

**Table S2. Age-matching exceptions for ADOS-2 comparators.**

| 3q29del proband age range (years) | Matched comparator exception | Group |
| --- | --- | --- |
| 0-5 | 1 month younger than criteria | 3q29del-ASD |
| 6-10 | 19 months younger than criteria | 3q29del-ASD |
| 6-10 | 15 months younger than criteria | 3q29del-ASD |
| 10-15 | 2 months younger than criteria | 3q29del-ASD |
| 10-15 | 5 months younger than criteria; 1 month older than criteria | 3q29del-ASD |
| 30-35 | 4 months younger than criteria | 3q29del+ASD |
| 36-40 | 3 months younger than criteria | 3q29del-ASD |

Exceptions to age matching for comparators in ADOS-2 data set.

**Table S3. First single word and first two-word phrase model results.**

| Comparison | ADI-R Communication domain | | ADI-R Nonverbal Communication sub-domain | |
| --- | --- | --- | --- | --- |
|  | Estimate | P value | Estimate | P value |
| First single word, full data set | 0.058 | 0.43 | 0.074 | 0.03 |
| First single word, 3q29del+ASD only | -0.159 | 0.45 | -0.047 | 0.50 |
| First single word, nsASD only | 0.141 | 0.08 | 0.157 | 0.005 |
| First two-word phrase, full data set | 0.076 | 0.07 | 0.039 | 0.06 |
| First two-word phrase, 3q29del+ASD only | 0.098 | 0.32 | 0.017 | 0.47 |
| First two-word phrase, nsASD only | 0.066 | 0.22 | 0.110 | 0.01 |

Summary of analyses of the relationship age at first single word and age at first two-word phrase with ADI-R domain B and ADI-R sub-domain B1.

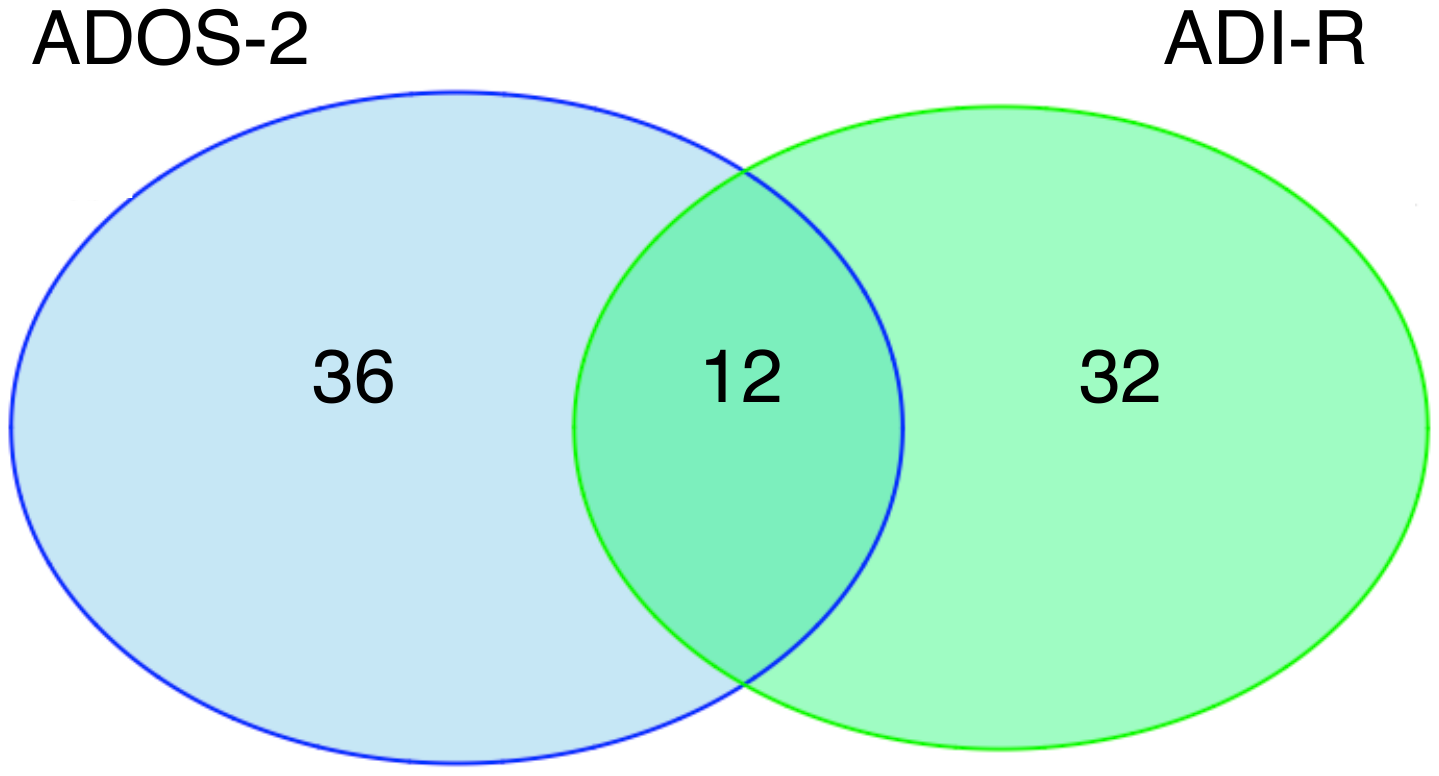

**Figures S1. Overlap in nsASD comparators between ADOS-2 and ADI-R.** Venn diagram showing the overlap in nsASD comparators ascertained from NDAR between the ADOS-2 and ADI-R data sets.
